# Automatic Bolus Tracking in Abdominal CT scans with Convolutional Neural Networks

**DOI:** 10.1101/2022.06.29.22276968

**Authors:** Angela Li, Peter B. Noël, Nadav Shapira

## Abstract

**Background:** Bolus tracking can optimize the time delay between contrast injection and diagnostic scan initiation in contrast-enhanced computed tomography (CT), yet the procedure is time-consuming and subject to inter- and intra-operator variances which affect the enhancement levels in diagnostic scans. The objective of the current study is to use artificial intelligence algorithms to fully automate the bolus tracking procedure in contrast-enhanced abdominal CT exams for improved standardization and diagnostic accuracy while providing a simplified imaging workflow.

**Methods:** This retrospective study used abdominal CT exams collected under a dedicated IRB. Input data consisted of CT topograms and images with high heterogeneity in terms of anatomy, sex, cancer pathologies, and imaging artifacts acquired with four different CT scanner models. Our method consisted of two sequential steps: (i) automatic locator scan positioning on topograms, and (ii) automatic ROI positioning within the aorta on locator scans. The task of locator scan positioning is formulated as a regression problem, where the limited amount of annotated data is circumvented using transfer learning. The task of ROI positioning is formulated as a segmentation problem.

**Results:** Our locator scan positioning network offered improved positional consistency compared to a high degree of variance in manual slice positionings, verifying inter-operator variance as a significant source of error. When trained using expert-user ground truth labels, the locator scan positioning network achieved a sub-centimeter error (9.76 ± 6.78 mm) on a test dataset. The ROI segmentation network achieved a sub-millimeter absolute error (0.99 ± 0.66 mm) on a test dataset.

**Conclusions:** Locator scan positioning networks offer improved positional consistency compared to manual slice positionings and verified inter-operator variance as an important source of error. By significantly reducing operator-related decisions, this method opens opportunities to standardize and simplify the workflow of bolus tracking procedures for contrast-enhanced CT.

## 1. Introduction

With advances in imaging technology and the exponential growth in the volume of medical imaging data, the current computed tomography (CT) workflow involves performing complex scanning protocols and processing requirements on a regular basis [1]. Manual procedures are not only time-consuming but also subject to inter-operator variances, creating potential for diagnostic inaccuracies [2]. In the new era of radiomics and deep learning, convolutional neural networks (CNNs) can serve as a highly effective tool for automating classification and detection tasks for medical imaging data [3,4].

Accurate enhancement phase determination is crucial for confident lesion characterization in contrast-enhanced CT, such as for assessing hypervascular hepatocellular carcinoma (HCC) during the arterial phase [5]. However, patient-specific variations such as heart rate, body weight, and circulation impairments can influence enhancement timing, introducing uncertainties and compromising confidence in diagnostic biomarkers [6,7]. Bolus tracking can help individualize time delays between contrast injection and diagnostic scan initiations by tracking the enhancement of radio-opaque contrast media (typically iodine) at a predefined operator-selected region [2,8]. The current bolus tracking workflow involves manual selection of a position along the patient axis for the locator scan on a two-dimensional (2D) CT topogram (also known as surview or scout scans), followed by manual selection of a region of interest (ROI) on the locator scan for tracking enhancement levels. Following the injection of intravenous contrast agent, low-dose tracker scans are executed continuously, e.g., every second, to monitor the increase in Hounsfield units (HU) within the selected ROI. Once a predefined HU-threshold is reached within the ROI, diagnostic scans are automatically initiated after predetermined time intervals that were optimized for the specific exam protocol or clinical indication [2,8].

Although tracking a small bolus of contrast media before diagnostic scans can help individualize the time delay, the technique is highly time-consuming [9]. Moreover, the manual selection procedures in the current bolus tracking workflow are prone to errors and intra-and inter-operator variance, particularly during the locator scan positioning step. To standardize the diagnosis process both between different patient populations and between different time-points during evaluations of a single patient [10], e.g., for treatment response assessment [11], several advancements in contrast-enhanced CT capabilities are required, including accurate contrast media quantification [12,13], normalization of iodine perfusion ratios [14], and more advanced fully automated bolus tracking techniques such as the one presented in this work.

The objective of the current study is to fully automate the bolus tracking procedure in contrast-enhanced abdominal CT exams using CNNs. Our method consisted of two sequential steps: (i) automatic locator scan positioning on topograms, and (ii) automatic ROI positioning within the aorta on locator scans. By reducing CT operator-related decisions, our method enables improved standardization and diagnostic accuracy, as well as greater efficiency in the diagnostic review process and clinical workflow. We present the following article in accordance with the TRIPOD reporting checklist.

## 2. Methods

In this study, we apply machine learning approaches using clinically available retrospective datasets to automate the bolus tracking procedure. Our method consists of two consecutive steps, each of which was developed and tested independently: (i) automatic determination of locator scan position on topograms, and (ii) automatic ROI positioning within the of the aorta on locator scans. A schematic of this two-step process is shown in Figure 1.

**Figure 1.**
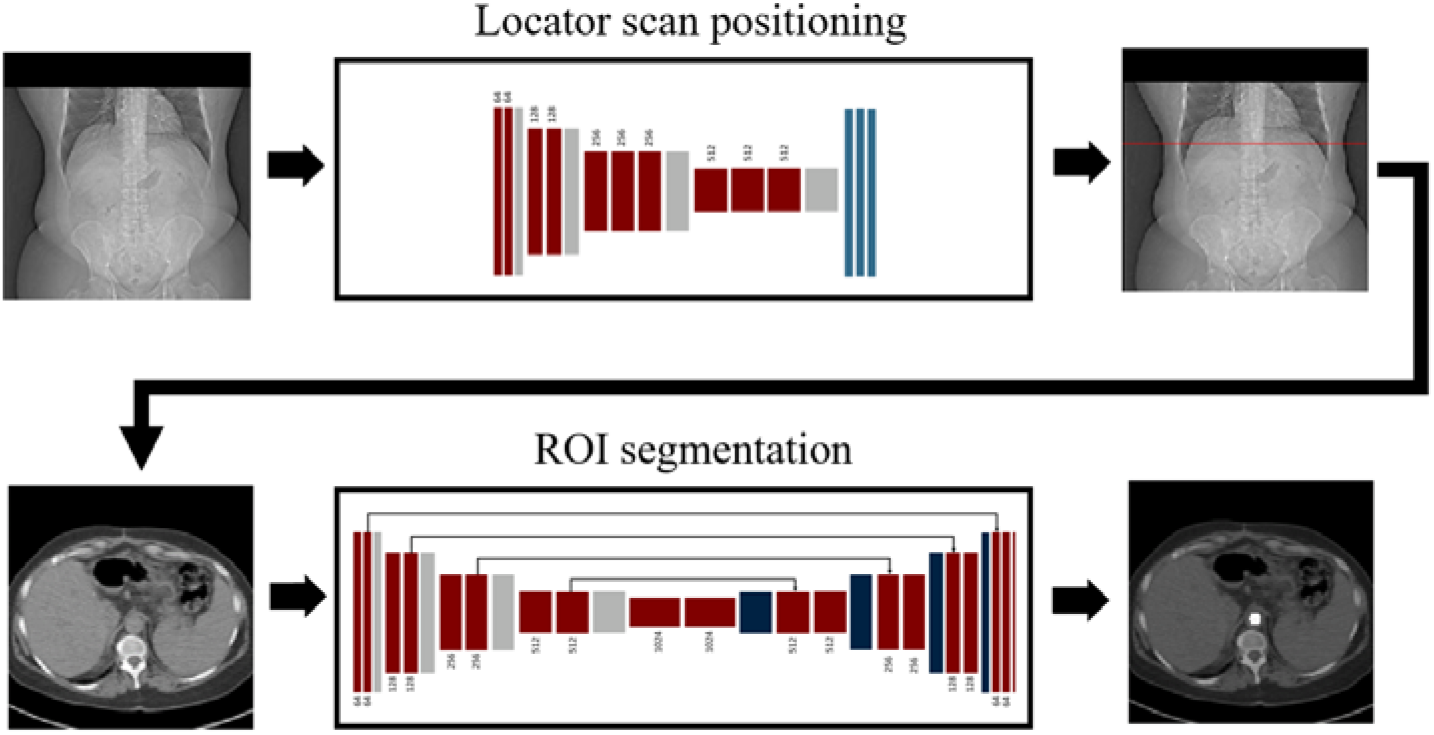
A two-step network pipeline for automatic bolus tracking. The method consisted of two sequential steps: (i) automatic locator scan positioning on topograms, and (ii) automatic ROI positioning within the aorta on locator scans. The task of locator scan positioning is formulated as a regression problem, where the limited amount of annotated data is circumvented using transfer learning. The task of aorta positioning is formulated as a segmentation problem.

While the segmentation task has been extensively addressed in the literature among the medical imaging community [15], methods on the automatic selection of a specific slice in an acquired CT topogram are extremely limited [16]. In recent years, researchers have begun to address this issue by proposing deep learning methods for the automatic detection of the third lumbar vertebra (L3) for body composition analysis. Such methods can be classified into the regression paradigm, which involves directly estimating the slice position given the entire CT scan in one-dimensional (1D) output [16] or 2D confidence maps [17], and the binary classification paradigm, which involves determining whether the target slice is present for each slice [18]. In this study, we retain the regression paradigm for our locator scan positioning task as it is more lightweight compared to binary classification methods which require extensive annotations of data.

### 2.1 Dataset collection and preparation

Retrospective topograms and locator scan images used in this study were collected from the Picture Archiving and Communication System (PACS) of our institute under a dedicated Institutional Review Board (IRB). Over three hundred CT examinations were identified and collected. Out of these cases, six were excluded due to locator scan positions that exceeded the length of the topogram. To study our algorithm’s generalizability, 10% of the 298 collected cases were randomly selected as the external validation (test) dataset (*N* = 30). Remaining data was randomly divided into training and validation sets in a ratio of 8:2 for development of our algorithm.

Collected CT exams showed a high level of heterogeneity in patient anatomy (sex, age, cancer pathology, and medical state) and consisted of four different scanner models, while topograms consisted of two different pixel spacing values (Table 1). Only coronal view topograms were retained for network training due to a small number of exams which included topograms in the sagittal view.

**Table 1.** Pixel spacing and scanner models of the collected datasets.

| Characteristic |  | Samples | % |
| --- | --- | --- | --- |
| Initial pixel spacing [mm/px] | 1 | 194 | 65 |
|  | 2 | 104 | 34 |
| Manufacturer’s Model Name | SOMATOM Force | 137 | 45 |
|  | SOMATOM Definition AS+ | 110 | 36 |
|  | SOMATOM Definition AS | 32 | 10 |
|  | SOMATOM Definition Flash | 14 | 4 |
|  | Other | 5 | 1 |

All coronal view topograms were resized to a 224×224 matrix size with a pixel spacing of 2.28 mm to ensure consistent input to the network. Ground-truth annotations of locator scan positions and ROI locations were extracted from the respective DICOM attributes ((0020,0032) Image Position Patient; (6000, 3000) Overlay Data) which record the geometric locations originally-selected by CT technologists. For both locator scan positioning and ROI positioning algorithms, the 2D matrix was duplicated in each color channel in order to match the required 3-channel input of the pre-trained models.

### 2.2 Locator scan positioning

In this study, the task of locator scan positioning is formulated as a regression problem, with the goal of predicting the slice position location along the patient axis (z-position) that corresponds to the anatomical location used for bolus tracking in these examinations. While the optimal locator scan position for bolus tracking is not well documented in the literature, for this work we adopted the general placement of 1 cm below the diaphragm proposed by Adibi *et al*. [2] as a proof of concept for this technique. Our algorithm can be easily retrained to target other positions or anatomies for the locator scan given the corresponding annotations.

Since modern machine learning algorithms require vast amounts of training data to achieve high accuracy, we adopted a transfer learning approach to improve sample efficiency [19]. In this framework, the weights of the network layers are initialized with the weights of a pre-trained CNN and frozen or fine-tuned to fit the target application. The networks were pre-trained on ImageNet, a classification database which contains over 14 million non-medical images separated into 1000 categories [20]. A randomly initialized fully connected layer with trainable parameters was stacked to the convolutional base to predict the locator scan position as an integer corresponding to the row number (in image pixels).

The mean squared error loss function was selected for training the network and the weights were optimized using Adam optimizer. We compared the performance of VGG16 [21], VGG19 [21], and ResNet50 [22] architectures to account for the effect of the number of trainable parameters and network depth on feature extraction. We also investigated the effect of introducing additional fully connected layers on algorithmic performance.

### 2.3 ROI segmentation

The task of ROI positioning was formulated as a segmentation problem, with the goal of localizing the aorta on locator scans. Input data consisted of 2D matrices of locator scans, with respective 2D binary masks indicating aorta position manually selected by the original CT technologist that serve as ground-truth segmentation targets. The binary masks were generated by selecting for and filling in the circle on the original overlay data extracted from the DICOMs (Figure 2).

**Figure 2.**
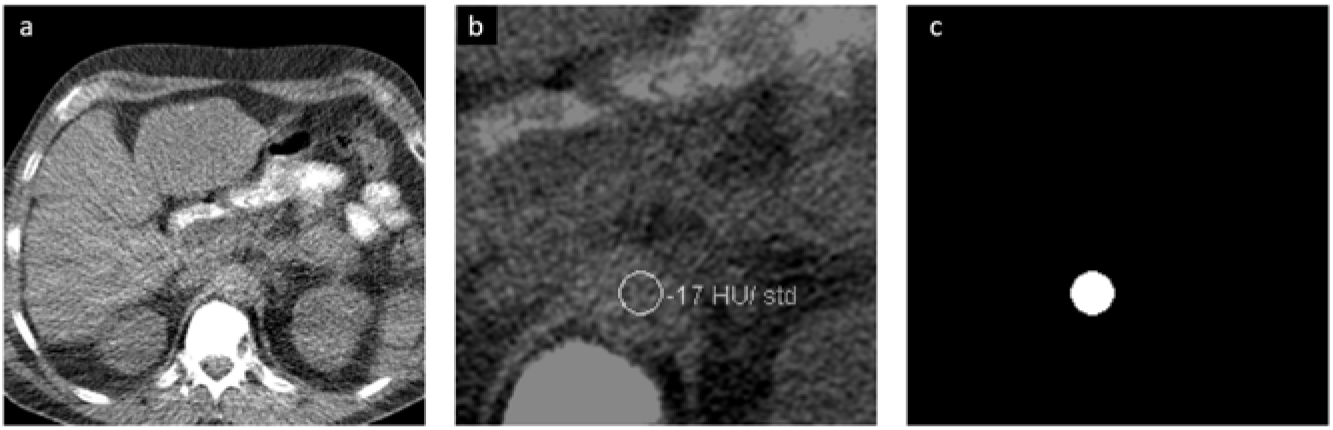
Example of generating the ground-truth inputs for the segmentation network, showing (a) original locator scan, (b) enlarged view of the aorta overlayed with binary mask extracted from DICOM data, and (c) processed mask indicating the aorta position that serves as ground-truth for the segmentation network.

A UNet was used to perform the segmentation task (Figure 3). The architecture consists of a multiple down-sampling and up-sampling blocks, each with a set of convolutional units, where each unit consists of a sequence convolution, batch normalization, and non-linear activation (ReLU) [23]. The loss function was selected as the binary cross-entropy and the model was optimized using Adam optimizer. Since the network generates 2D matrices as output, no further adjustments were made to the original architecture. Network predictions consisted of 2D binary masks displaying ROI location predictions. Error was quantified by the Euclidean distance between the center of mass 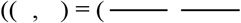 of the expected and predicted ROI in millimeters.

**Figure 3.**
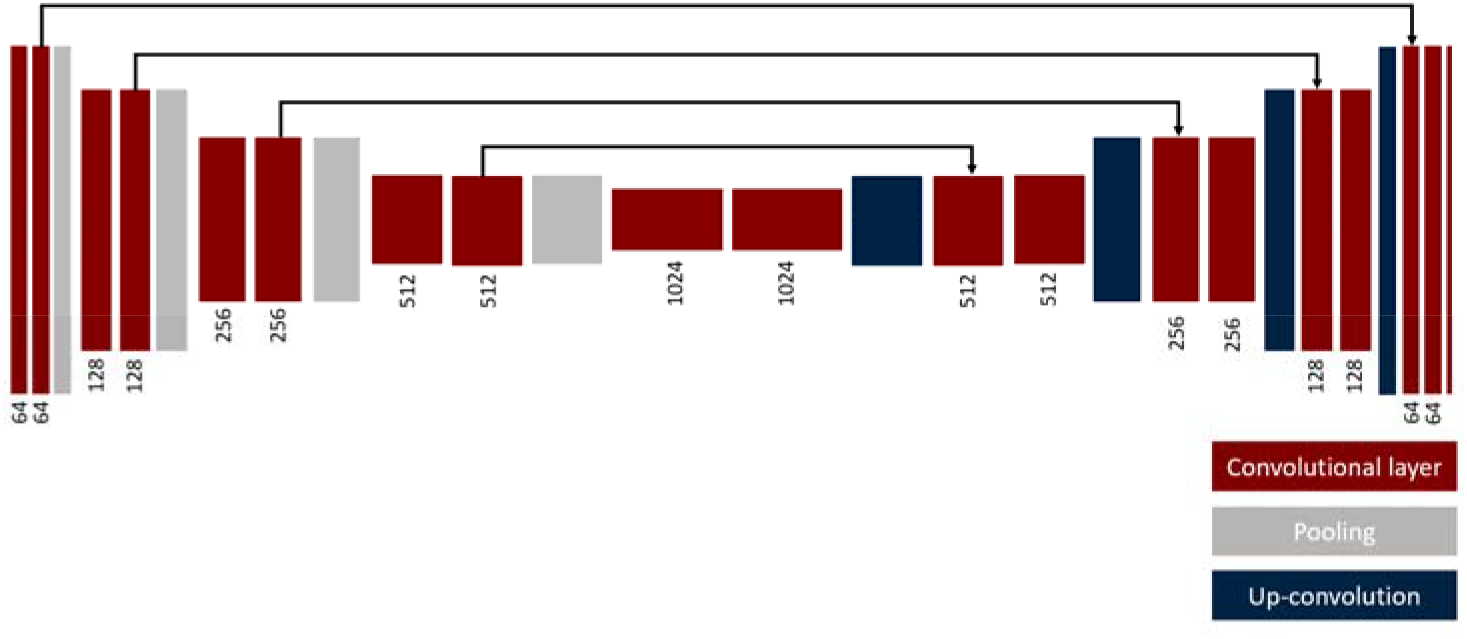
The UNet architecture consists of a multiple down-sampling (left) and up-sampling blocks (right). Each block consists of a set of convolutional units, where each unit consists of a sequence convolution, batch normalization, and non-linear activation (ReLU).

## 3. Results

### 3.1 Automatic locator scan positioning

Performance of the locator scan positioning model was investigated by computing the prediction error on the external test dataset (*N* = 30). The prediction error for a single CT scan is computed as the absolute difference between the network prediction and target in millimeters. We report the mean (μ) and the standard deviation of the prediction error (σ) over the entire test set in Table 2.

**Table 2:**
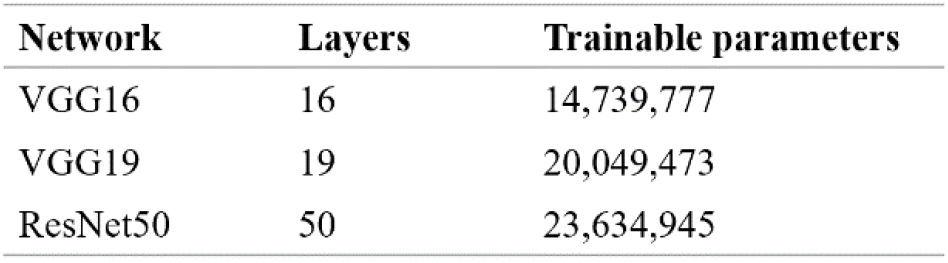
Network architectures used in this study. The architectures varied in network depth and number of trainable parameters.

**Table 2.**
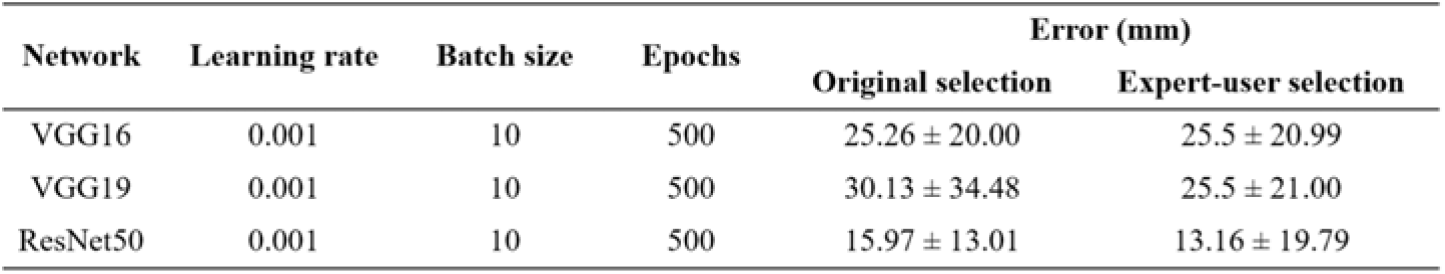
Comparison of locator scan positioning algorithms. Performance was evaluated by computing the prediction error on a test dataset consisting of 30 unseen topograms.

To account for the possible impact of intra-operator variance, we also trained and tested the models using expert-level labels of a single user in addition to those originally extracted from the DICOM attributes. The expert-level ground truth labels were retrospectively selected by Penn Medicine’s Lead CT Technologist with the aid of a dedicated graphical user interface (GUI) that we implemented where the annotator clicked on the location of the locator scan for each CT topogram and the selected position along the patient axis was recorded. No reference landmarks, such as the original locator scan position, were provided to the annotator.

Of the three models, the ResNet50-based model yielded the smallest predictive error (13.16 ± 19.79 mm). We found that adding additional fully connected layers at the end of the network did not improve algorithmic performance. Importantly, models that were trained on a single expert-level operator labels yielded a smaller or similar predictive error, verifying inter-operator variance as a significant source of error. An example of the inter-operator variance of the original selections can be clearly observed when compared to the respective selections made by a single expert-level operator (Figure 4). The network predictions provide improved positional consistency compared to the high degree of variance in manual slice positionings performed by CT operators, particularly when examined relative to distinct anatomical structures such as the diaphragm.

**Figure 4.**
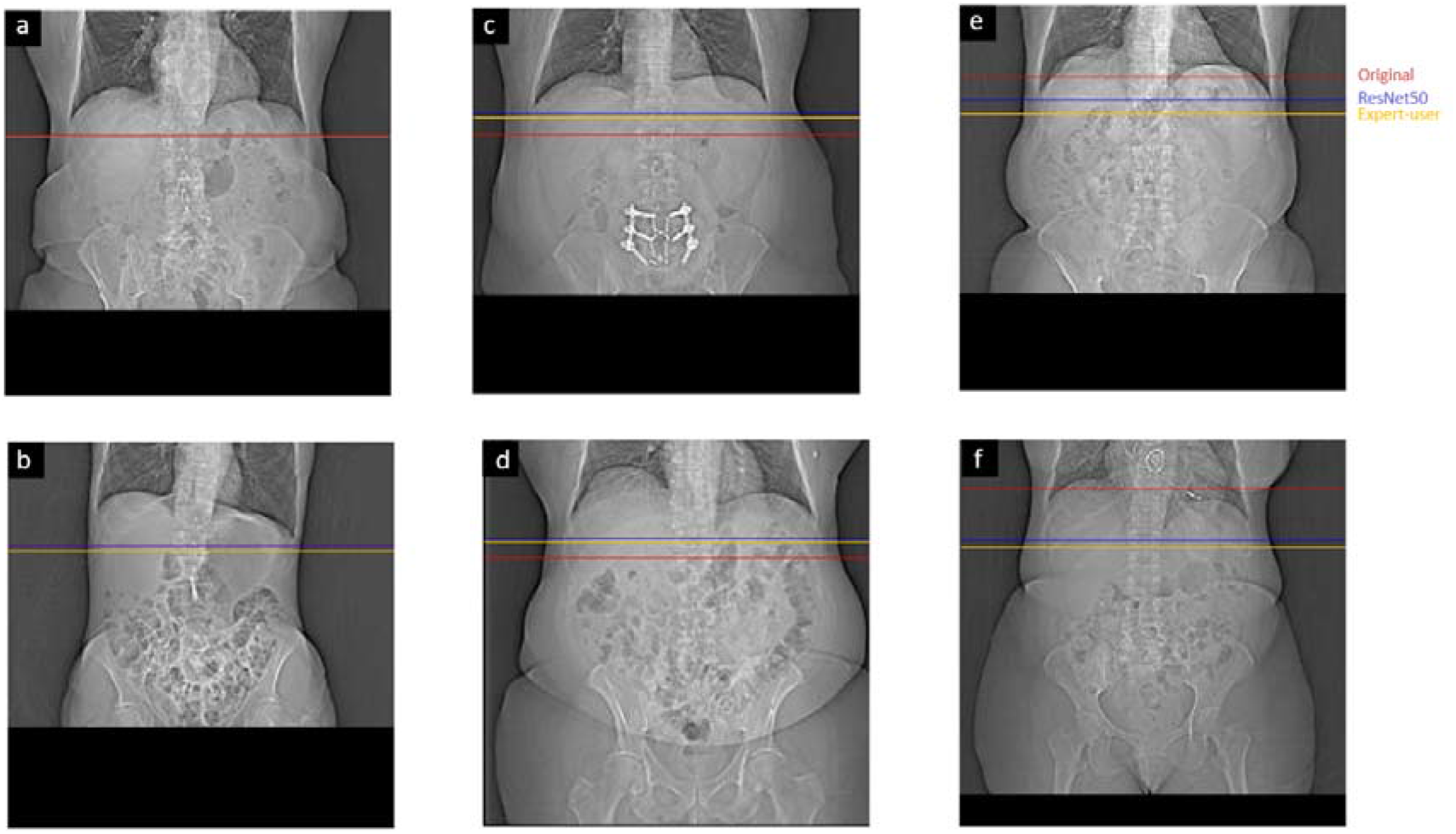
Example coronal topograms of a test dataset showing locator scan positions selected by original operator (red), single expert-level user (yellow), and ResNet50-based model (blue). Examples show both good agreement (a,b) and large error (c-f). We attribute a significant portion of the network’s error to observed large inter-operator variance.

To better understand the high standard deviation in our model’s predictive error, we computed the error distribution of all training and test samples (Figure 5). Good agreement is observed across the majority of samples, with 90% of samples having errors below a single centimeter. Examples of coronal topograms that resulted in good agreement with the locations selected by the expert-level user are shown in Figure 6. While the optimal locator scan position for bolus tracking is not well documented in the literature, Adibi *et al*. have proposed a general placement of 1 cm below the diaphragm [2], consistent with our CNN predictions.

**Figure 5.**
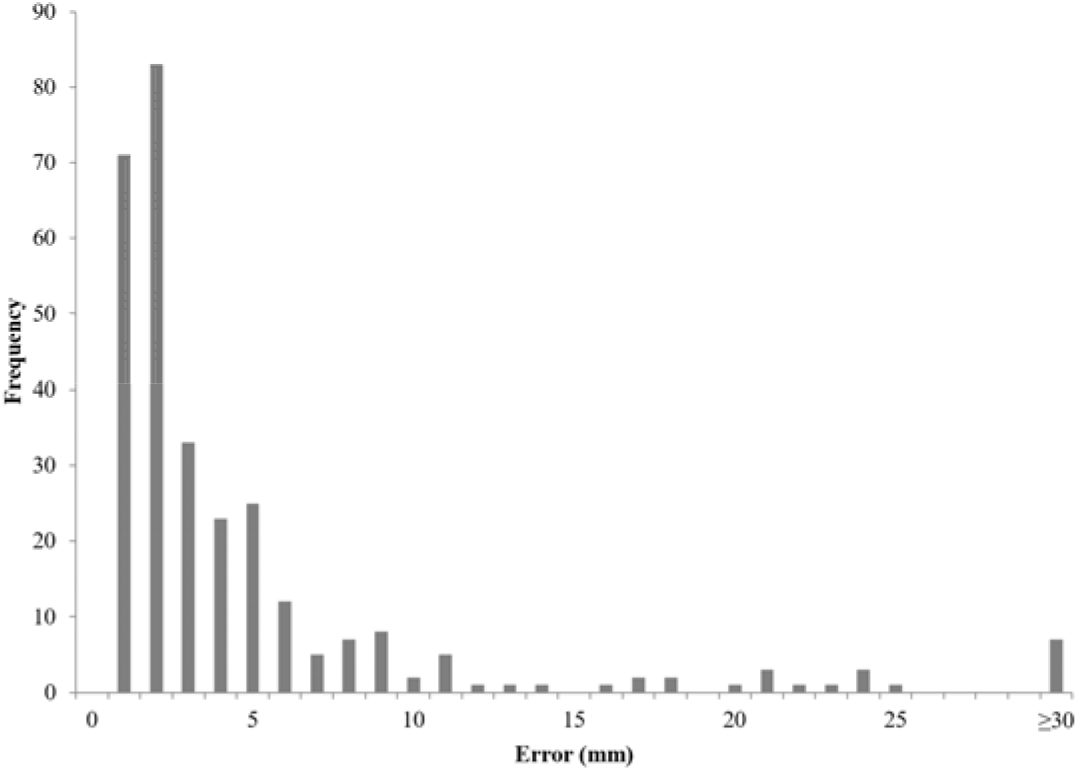
Distribution of absolute error of across all training and test samples of a ResNet50 model trained using expert-level labels.

**Figure 6.**
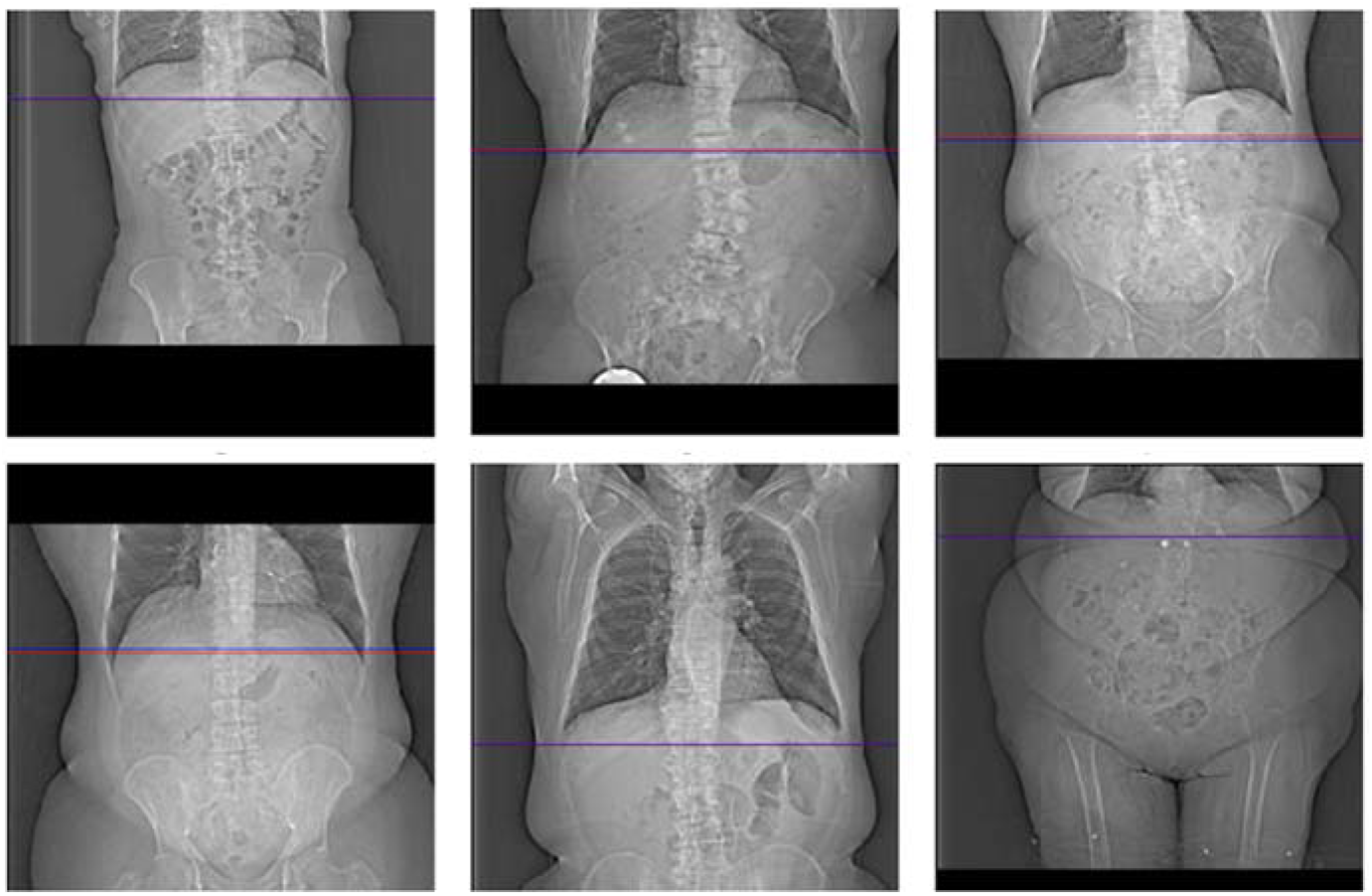
Example coronal topograms of in the training dataset showing locator scan positions selected by a single expert-level user (red), and CNN (blue). Good agreement is observed across the majority (90%) of samples.

We subsequently analyzed samples with large error (here defined as those with predictive error ≥30 mm). The network showed a large error on a total of 7 out of 298 samples (Figure 7). These errors can be classified into two categories: prediction errors and labelling errors. For the first category, the network predictions were approximately a single vertebra away from that selected location by the operator (Figure 7a-7c). Labelling errors were observed in samples in which there was a clear inconsistency in the operator’s ground-truth labeling (Figure 7d-7g). However, the network was able to accurately predict locator scan positions in these cases, even in the presence of artifacts and implants. After excluding the four samples containing labelling errors, our model yielded a final average predictive error of 9.76 ± 6.78 mm.

**Figure 7.**
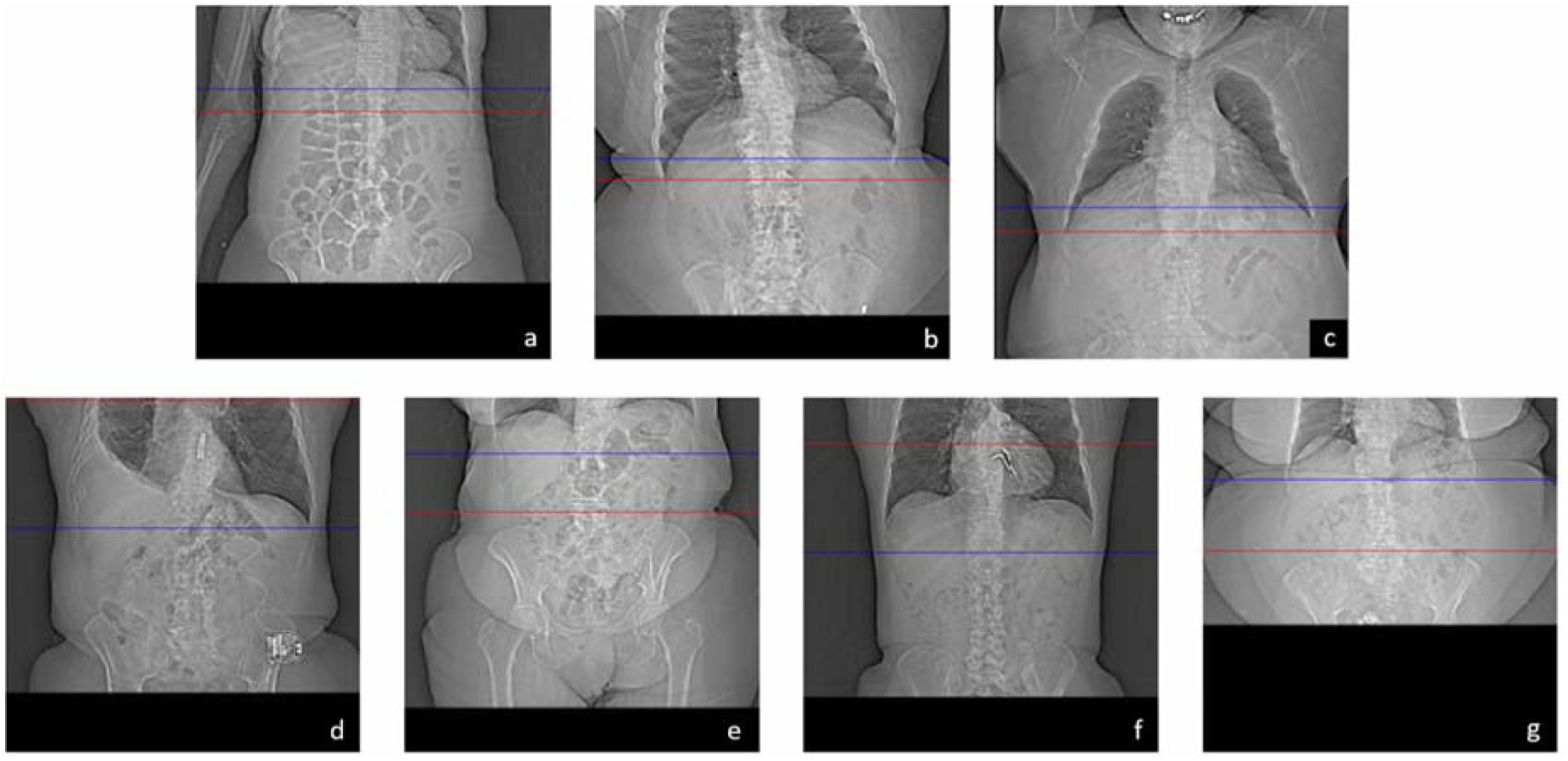
Training samples with large error showing locator scan positions selected by the expert-level user (red), and CNN (blue). The network prediction showed an error greater than 30 mm on a total of 7 out of 298 samples. These errors can be divided into two categories: prediction errors (a-c) and labelling errors (d-g). Importantly, the network was able to accurately predict locator scan positions in the presence of artifacts and implants (d, g).

### 3.2 Automatic ROI positioning

Performance of the segmentation network was investigated by computing the Euclidean distance between the center of mass of the expected and predicted ROI in millimeters. The raw output of the ROI positioning network is a 2D confidence map with each pixel value in the range (0,1). To generate a binary predicted mask used for evaluation, we applied a binary threshold of value of 0.9999999, under which each pixel of the mask is set to either 0 or 1 depending on its intensity value relative to the threshold (Figure 8).

**Figure 8.**
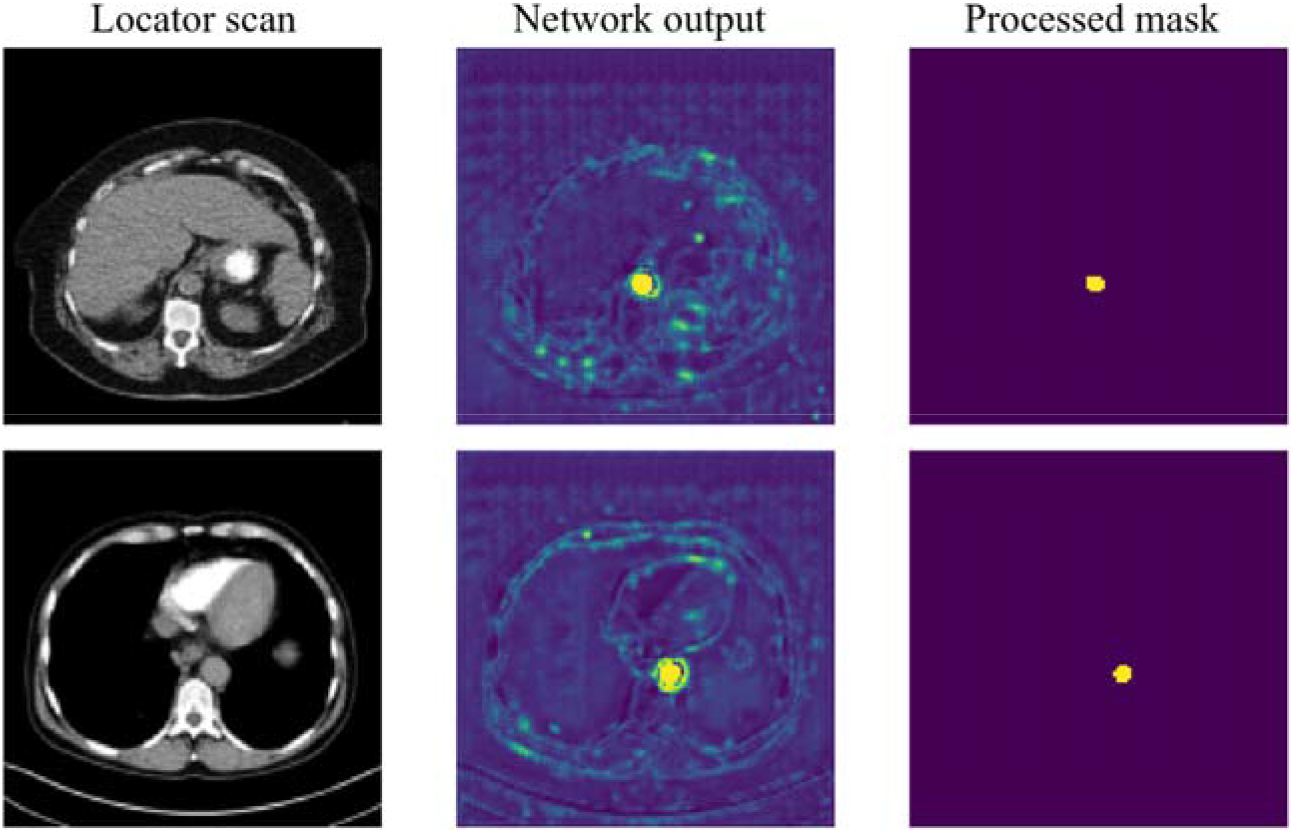
Examples of binary thresholding to produce the final predicted binary mask used for evaluation.

Overall, our ROI segmentation network showed no significant difference with the original manual annotations and achieved a sub-millimeter error on the test dataset (0.99 ± 0.66 mm). Furthermore, the network accurately predicted ROI position in the presence of imaging artifacts, validating the network’s robustness (Figure 9).

**Figure 9.**
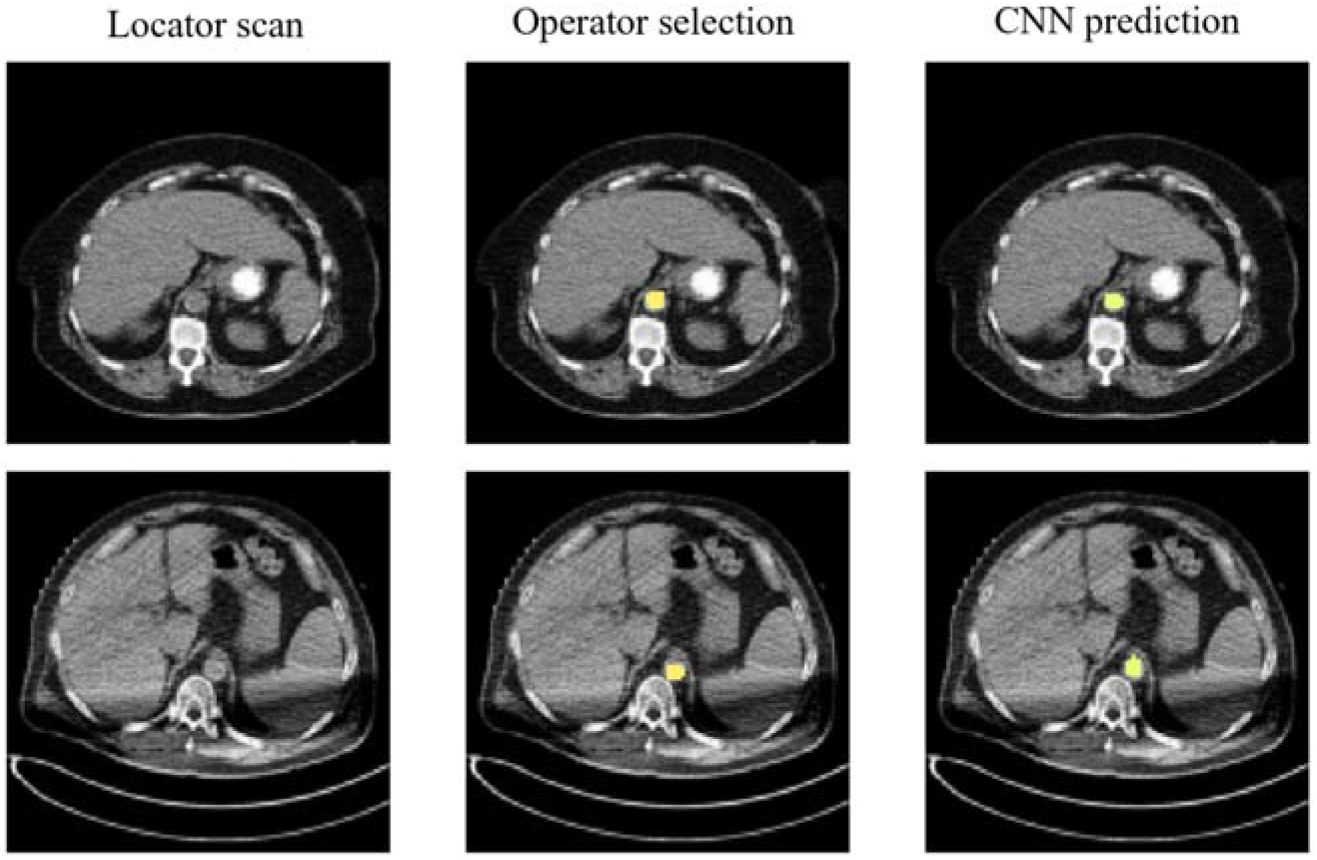
Examples of locator scans with respective expected and predicted binary masks on a test dataset. Our ROI segmentation network showed no significant difference with the original manual annotations and was able to accurately segment the ROI in the presence of imaging artifacts (bottom row).

## 4 Discussion

The current computed tomography (CT) workflow involves performing complex scanning protocols and processing requirements on a regular basis [1]. Computer-assisted bolus tracking can optimize time delays between contrast injection and diagnostic scan initiation in contrast-enhanced CT compared to fixed delay techniques; however, the procedure is time-consuming and subject to intra- and inter-operator variances which may affect enhancement levels in diagnostic scans [9]. In the present study, we developed machine learning algorithms to automate and standardize the (i) locator scan positioning and (ii) aorta positioning tasks of the bolus tracking procedure. The task of locator scan positioning is formulated as a regression problem, where the limited amount of annotated data was circumvented using transfer learning, while the task of ROI segmentation is formulated as a segmentation problem. The CNNs were trained using retrospective clinical datasets (*N* = 298), and evaluated on an unseen test dataset (*N* = 30).

Our locator scan positioning network offered greater positional consistency compared to manual slice positionings, verifying intra- and inter-operator variance as major sources of error in the bolus-tracking procedure. When trained using single-user ground truth labels, the locator scan positioning network achieved a 9.76 ± 6.78 mm error on a test dataset. Our ROI segmentation network showed no significant difference with manual annotations and achieved a sub-millimeter error between the predicted and expected ROI (0.99 ± 0.66 mm). This automated pipeline offers improved standardization and accuracy in the bolus tracking procedure compared to manual methods, highlighting the potential for greater consistency between examinations of different patients as well as between examinations of a single patient at different evaluation time points.

Significant reductions in inter-operator variance open opportunities to evaluate, model, and reduce the effects of individualized patient kinetics, allowing for further refinement of diagnostic CT acquisitions. While greater accuracy in the bolus-tracking procedure increases the likelihood of reaching optimal phase timings, combining this pipeline with automated phase-identification algorithms can ensure that the optimal phase is successfully reached in a patient, regardless of anthropomorphic variation and hemodynamic status [24,25]. Importantly, non-optimal portal venous phase acquisition timing occurs in one out of three patients in multicenter clinical trials, significantly altering tumor density measurements [26]. The ability to accurately identify phase timings also allows for further refinement of diagnostic CT procedures, through standardizing iodine injection and uptake, and potentially reducing iodine dose. Finally, this pipeline may be useful tool for the standardization of novel dual-contrast imaging procedures, in which information of the first contrast is essential to accurately forecast the time point of maximal arterial enhancement by the second contrast agent [27].

There are limitations of our study. First, inter- and intra-operator variances contributed to a lack of accurate ground-truth labels for the locator scan positioning network. Revision of ground-truth labels, such as by taking the average of multiple CT technologist annotations [17], may provide more accurate training data for improved network performance. Second, the present study does not address patient outcomes, making it difficult to assess the effect of slice localization offsets and segmentation differences on the final stratification of patients. Moving forward, clinical trials will effectively determine the advantages of the automatic selection suggestions and its incorporation in larger studies that aim to advance CT evaluations.

In conclusion, we developed a machine learning pipeline to automate and standardize the locator scan positioning and ROI segmentation tasks in the bolus tracking workflow. By significantly reducing operator-related decisions, this method opens opportunities to evaluate, model, and reduce the effect of patient kinetics in contrast-enhanced CT exams.

## Data Availability

All data produced in the present work are contained in the manuscript.

## 5 Acknowledgements

We acknowledge support through the National Institutes of Health (R01EB030494).

Special thanks to Penn Medicine’s Lead CT Technologist, Michael Colfer, for expert-level labeling of the locator scan positionings.

## Footnote

### Reporting Checklist

The authors have completed the TRIPOD checklist.

### Conflicts of Interest

All authors have completed the ICMJE uniform disclosure form. PN receives research grant funding from Philips Healthcare and Siemens Medical Solutions USA. The other authors have no conflicts of interest to declare.

### Ethical Statement

The authors are accountable for all aspects of the work in ensuring that questions related to the accuracy or integrity of any part of the work are appropriately investigated and resolved. The study was conducted in accordance with the Declaration of Helsinki (as revised in 2013). The study was approved by the institutional ethics board of the University of Pennsylvania and individual consent for this retrospective analysis was waived.

### Author contributions

(I) Conception and design: P Noël, N Shapira; (II) Administrative support: P Noël, N Shapira; (III) Provision of study materials or patients: P Noël, N Shapira; (IV) Collection and assembly of data: A Li; (V) Data analysis and interpretation: A Li, N Shapira; (VI) Manuscript writing: All authors; (VII) Final approval of manuscript: All authors.

## Notes

### Funding Statement

This study did not receive any funding.

### Author Declarations

IRB of University of Pennsylvania gave ethical approval for this work

## References

[1] Cody DD, Dillon CM, Fisher TS, Liu X, McNitt MF, Patel V. AAPM Medical Physics Practice Guideline 1.b: CT protocol management and review practice guideline. :7.

[2] Adibi A, Shahbazi A. Automatic Bolus Tracking Versus Fixed Time-Delay Technique in Biphasic Multidetector Computed Tomography of the Abdomen. Iran J Radiol [Internet]. 2014 Jan 30; 10(4). Available from: https://brief.land/iranjradiol/articles/72137.html

[3] Zreik M, van Hamersvelt RW, Wolterink JM, Leiner T, Viergever MA, Isgum I. A Recurrent CNN for Automatic Detection and Classification of Coronary Artery Plaque and Stenosis in Coronary CT Angiography. IEEE Trans Med Imaging. 2019 Jul;38(7):1588–98.

[4] Irmak E. Multi-Classification of Brain Tumor MRI Images Using Deep Convolutional Neural Network with Fully Optimized Framework. Iran J Sci Technol Trans Electr Eng. 2021 Sep;45(3):1015–36.

[5] Lee JH, Lee JM, Kim SJ, Baek JH, Yun SH, Kim KW, et al. Enhancement patterns of hepatocellular carcinomas on multiphasic multidetector row CT: comparison with pathological differentiation. Br J Radiol. 2012 Sep;85(1017):e573–83.

[6] Bae KT, Seeck BA, Hildebolt CF, Tao C, Zhu F, Kanematsu M, et al. Contrast enhancement in cardiovascular MDCT: effect of body weight, height, body surface area, body mass index, and obesity. AJR Am J Roentgenol. 2008;190(3):777–84.

[7] Bae KT, Tao C, Gurel S, Hong C, Zhu F, Gebke TA, et al. Effect of patient weight and scanning duration on contrast enhancement during pulmonary multidetector CT angiography. Radiology. 2007;242(2):582–9.

[8] Fukukura Y, Takumi K, Kamiyama T, Shindo T, Higashi R, Nakajo M. Pancreatic adenocarcinoma: a comparison of automatic bolus tracking and empirical scan delay. Abdom Imaging. 2010 Oct;35(5):548–55.

[9] Ramos-Duran LR, Kalafut JF, Hanley M, Schoepf UJ. Current Contrast Media Delivery Strategies for Cardiac and Pulmonary Multidetector-row Computed Tomography Angiography. Journal of Thoracic Imaging. 2010 Nov;25(4):270–7.

[10] McNitt-Gray MF, Bidaut LM, Armato SG, Meyer CR, Gavrielides MA, Fenimore C, McLennan G, Petrick N, Zhao B, Reeves AP, Beichel R, Kim HJ, Kinnard L. Computed tomography assessment of response to therapy: tumor volume change measurement, truth data, and error. Transl Oncol. 2009 Dec;2(4):216–22. doi: 10.1593/tlo.09226. PMID: 19956381; PMCID: PMC2781084.

[11] Sheikhbahaei S, Mena E, Yanamadala A, Reddy S, Solnes LB, Wachsmann J, et al. The Value of FDG PET/CT in Treatment Response Assessment, Follow-Up, and Surveillance of Lung Cancer. American Journal of Roentgenology. 2017 Feb;208(2):420–33.

[12] Hua C ho, Shapira N, Merchant TE, Klahr P, Yagil Y. Accuracy of electron density, effective atomic number, and iodine concentration determination with a dual-layer dual-energy computed tomography system. Med Phys. 2018 Jun;45(6):2486–97.

[13] Sellerer T, Noël PB, Patino M, Parakh A, Ehn S, Zeiter S, et al. Dual-energy CT: a phantom comparison of different platforms for abdominal imaging. Eur Radiol. 2018 Jul;28(7):2745–55.

[14] Zopfs D, Graffe J, Reimer RP, Schäfer S, Persigehl T, Maintz D, et al. Quantitative distribution of iodinated contrast media in body computed tomography: data from a large reference cohort. Eur Radiol. 2021 Apr;31(4):2340–8.

[15] Pham DL, Xu C, Prince JL. Current Methods in Medical Image Segmentation. Annu Rev Biomed Eng. 2000 Aug;2(1):315–37.

[16] Belharbi S, Chatelain C, Hérault R, Adam S, Thureau S, Chastan M, Modzelewski R. Spotting L3 slice in CT scans using deep convolutional network and transfer learning. Comput Biol Med. 2017 Aug 1;87:95–103. doi: 10.1016/j.compbiomed.2017.05.018. Epub 2017 May 19. PMID: 28558319.

[17] Kanavati F, Islam S, Aboagye EO, Rockall A. Automatic L3 slice detection in 3D CT images using fully-convolutional networks. 181109244 [cs] [Internet]. 2018 Nov 22 [cited 2022 Mar 23]; Available from: http://arxiv.org/abs/1811.09244

[18] Dabiri S, Popuri K, Ma C, Chow V, Feliciano EMC, Caan BJ, et al. Deep learning method for localization and segmentation of abdominal CT. Computerized Medical Imaging and Graphics. 2020 Oct 1;85:101776.

[19] Weiss K, Khoshgoftaar TM, Wang D. A survey of transfer learning. J Big Data. 2016 Dec;3(1):9.

[20] Deng J, Dong W, Socher R, Li L-J, Kai Li, Li Fei-Fei. ImageNet: A large-scale hierarchical image database. In: 2009 IEEE Conference on Computer Vision and Pattern Recognition [Internet]. Miami, FL: IEEE; 2009 [cited 2022 Mar 22]. p. 248–55. Available from: https://ieeexplore.ieee.org/document/5206848/

[21] Simonyan K, Zisserman A. Very Deep Convolutional Networks for Large-Scale Image Recognition. 14091556 [cs] [Internet]. 2015 Apr 10 [cited 2022 Mar 22]; Available from: http://arxiv.org/abs/1409.1556

[22] He K, Zhang X, Ren S, Sun J. Deep Residual Learning for Image Recognition. 151203385 [cs] [Internet]. 2015 Dec 10 [cited 2021 Aug 10]; Available from: http://arxiv.org/abs/1512.03385

[23] Ronneberger O, Fischer P, Brox T. U-Net: Convolutional Networks for Biomedical Image Segmentation. 150504597 [cs] [Internet]. 2015 May 18 [cited 2022 Mar 22]; Available from: http://arxiv.org/abs/1505.04597

[24] Ma J, Dercle L, Lichtenstein P, Wang D, Chen A, Zhu J, et al. Automated Identification of Optimal Portal Venous Phase Timing with Convolutional Neural Networks. Academic Radiology. 2020 Feb;27(2):e10–8.

[25] Silverman PM, Brown B, Wray H, Fox SH, Cooper C, Roberts S, et al. Optimal contrast enhancement of the liver using helical (spiral) CT: value of SmartPrep. American Journal of Roentgenology. 1995 May;164(5):1169–71.

[26] Dercle L, Lu L, Lichtenstein P, Yang H, Wang D, Zhu J, et al. Impact of Variability in Portal Venous Phase Acquisition Timing in Tumor Density Measurement and Treatment Response Assessment: Metastatic Colorectal Cancer as a Paradigm. JCO Clinical Cancer Informatics. 2017 Nov;(1):1–8.

[27] Muenzel D, Daerr H, Proksa R, Fingerle AA, Kopp FK, Douek P, et al. Simultaneous dual-contrast multi-phase liver imaging using spectral photon-counting computed tomography: a proof-of-concept study. Eur Radiol Exp. 2017 Dec;1(1):25.

